# Graph Neural Network Modelling as a potentially effective Method for predicting and analyzing Procedures based on Patient Diagnoses

**DOI:** 10.1101/2021.11.25.21266465

**Authors:** Juan G. Diaz Ochoa, Faizan Mustafa

## Abstract

**Background:** Currently, the healthcare sector strives to increase the quality of patient management and improve the economic performance of healthcare providers. The data contained in electronic health records (EHRs) offer the potential to discover relevant patterns that aim to relate diseases and therapies, and thus discover patterns that could help identify empirical medical guidelines that reflect best practices in the healthcare system. Based on this pattern identification, it is then possible to implement recommendation systems based on the idea that a higher volume of procedures is associated with high-quality models.

**Methods:** Although there are several applications that use machine learning methods to identify these patterns, this identification is still a challenge, in part because these methods often ignore the basic structure of the population, considering the similarity of diagnoses and patient typology. To this end, we have developed graph methods that aim to cluster similar patients. In such models, patients are linked when the same or similar patterns can be observed for these patients, a concept that enables the construction of a network-like structure. This structure can then be analyzed with Graph Neural Networks (GNN) to identify relevant labels, in this case the appropriate medical procedures.

**Results:** We report the construction of a patient Graph structure based on basic patient’s information like age and gender as well as the diagnoses and trained GNNs models to identify the corresponding patient’s therapies using a synthetic patient database. We compared our GNN models against different baseline models (using the SCIKIT-learn library of python) and compared the performance of the different model methods. We have found that GNNs are superior, with an average improvement of the *f1* score of 6.48% respect to the baseline models. In addition, the GNNs are useful for performing additional clustering analyses that allow specific identification of specific therapeutic clusters related to a particular combination of diagnoses.

**Conclusions:** We found that GNNs are a promising way to model the distribution of diagnoses in a patient population and thus better model how similar patients can be identified based on the combination of morbidities and comorbidities. Nevertheless, network building is still challenging and prone to prejudice, as it depends on how ICD distribution affects the patient network embedding space. This network setup requires not only a high quality of the underlying diagnostic ecosystem, but also a good understanding of how to identify related patients by disease. For this reason, additional work is needed to improve and better standardize patient embedding in graph structures for future investigations and applications of services based on this technology, and therefore is not yet an interventional study.

## 1 Introduction

The analysis of electronic patient records (EHRs) is not only a source of information about the patient’s condition to obtain useful data on diagnosis and patient management^1^, but also to find empirical guidelines for the best possible medical procedures for the patient (Reddy and Aggarwal, 2015) (Malik et al., 2021). Therefore, the integration of the information contained in EHRs and their modelling is essential to improve patient management, for example for the design of individual and adaptive therapies (Diaz Ochoa and Weil, 2019), or the extraction of quality indicators. Up to the present such analysis are performed based on well-established parameters like Inpatient Quality Indicators (IQI), Prevention Quality Indicators (PQI) or Patient Safety Indicators (PSI) (Farquhar, 2008). However, these parameters are measured as consolidated metrics, ignoring inherent patterns and dynamics in the way therapies and medical procedures are assigned to patients.

In this work, we want to evaluate IQI essentially as a proof of concept, i.e. the evaluation of procedures where there is evidence that a higher volume of processes is associated with its high quality (Farquhar, 2008). To this end, we do not evaluate consolidated volume, but point patterns for individual patients with chronic disease to find out how diagnoses (encoded by the International Classification of Diseases ICD-10) are correlated with the medical procedures. In this way, we have developed a novel method based on Graph Neural Networks (GNNs), which enables a detailed and coarse-grained IQI analysis.

With regard to this problem, Hema et al. proposed graph-like structures and methods that *capture and represent these codes in the form of knowledge from semantic structured repository with better materialization, quality, and repository utilization, along with logical unity and logical entailments to form conceptual relationships between them* (Hema and Justus, 2015a). This method aligns towards linked health data, which is considered as a relevant way to leverage the complex information stored in the health system (Kotwal et al., 2016). This groundbreaking work therefore had the basic concepts for ICD clustering, especially using graph concepts.

In our case, we analyze data from patients with multiple morbidities, which means that each patient can be assigned specific combinations of ICD groups/clusters. Accordingly, groups of therapies and accountable medical procedures are coded by so called Therapy Keys (TKs). Good disease management requires a good knowledge of how ICD and TK clusters arise and correlate in patient populations. Of course, the detection of these clusters leads to improved clinical and economic management of the disease. But in order to get a better knowledge of the system, we face the following problems:

- Unknown frequency of use of TKs regarding ICDs. Discover clusters of ICDs and TKs
- Unknown correlations of TKs to ICDs in real databases
- Unknown patient clustering considering diseases and therapies

The hope is to have good methods to efficiently extract relevant features from electronic health records (EHR) with little information loss to solve these problems. *For example, the codification and representation of knowledge from structured and unstructured repositories was the prerequisite for* the design *of expert systems* based on the use of graph representations (Hema and Justus, 2015b).

There are already ongoing projects leading to the Graph representation of ICDs as well as GNNs to generalize the ability of learning implicit medical concept structures to a wide range of data sources^2^. These applications range from the evaluation of health-records for the prediction of the stay time of patients in intensive care stations (Rocheteau et al., 2021), the analysis of the ontology structure of ICD-9 codes (Rasmy et al., 2021; Shang et al., 2019), and the application of Graph-Concepts to the prediction of adverse pharmacological events by the modelling of the interactome and polypharmacy (Zitnik et al., 2018). Based on these first research works it is to expect that the in the next years the application of GNNs to the analysis of EHRs will become more ubiquitous, in particular for the empirical identification of medical guidelines.

Of course, medical guidelines are available from the literature and are currently applied in health service. The problem is the lack of information about how these guidelines are currently applied in real life. For this reason, methods are required for empirical research about how these guidelines “emerge” from the health records. Therefore, the main Goal of this work is essentially to identify correlations between TKs and ICD-10 groups, such that TKs can be predicted by providing the patient’s ICDs as well patient’s metadata. An additional feature will be the identification of patient clusters, as well as the characterization of certain ICD combination and structures, which will allow the identification of groups of patients with specific morbidity combinations. Based on the correlation between ICD and TKs it will be therefore possible to assess if such patient groups are getting the same therapy, or if there are deviations in such clusters.

While this goal has a practical character, i.e., the application of clustering methods to identify correlations and patient clusters, we also need to solve a methodological problem, namely the identification of the best possible method to reconstruct the relationship between similar patients.

In the first part, we provide an overview of the methodology applied in this study, including the description of the basic input structures, the description of the base line models, the method applied to the ICD-Graph definition and the implementation of the GNN method to predict TKs. In the results section we present the output of the analysis and make a critical comparison between the accuracy of the GNN model respect the baseline models, as well as the application of this methodology for the subsequent clustering analysis. Thereafter, we make a discussion of the present model and results, also considering potential ethical concerns. Finally, we present the main conclusions of this study.

## 2 Methods

### 2.1 Data extraction and synthetization

For this study we have obtained a fully synthetic high qualitatively database (with less than 1% mean error) computed from an anonymized database. The use of synthetic records is a *risk avoidance strategy for protecting patient information eliminating any potential risk of data privacy breaches suffered by de-identified PHI* ^*3*^ (Gallagher et al., 2018) (Phillips et al., 2017). Despite this synthetic data could bias real data contained in EHRs (Chen et al., 2021), we opted to implement this study using this kind of data since we basically aim to explore the performance of different algorithms before using them for medical decisions or for countability in medicine.

In addition, we cloned the synthetic patient information and thus generated a completely new and larger database (data augmentation) for machine learning porpoise. The data cloning was generated using the synthpop package implemented in R (Nowok et al., 2016). In table 1 we present the main parameters of this database.

**Table 1.** Principal parameters in the synthetic database

| Variable | Meaning | Kind |
| --- | --- | --- |
| GI | Geographical Identification | Character |
| ID | Internal Patient Identification | Numerical |
| Gender | - | Binary |
| Age | - | Numerical |
| TK | Therapy Key, equivalent to medical procedures | Character |
| ICD | International Classification of Diseases | Character |

### 2.2 Multilabel matrices

The task is to predict the Therapy Keys (TK) for a patient depending on the diagnosis (ICDs) and additional metadata, like patient’s age, gender, and pertinence to a specific health center (geographical distribution). To this end we represent the TKs and ICDs as a multi-label matrix 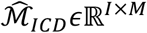, where *M* is the number of unique ICDs and *I* is the total number of patients, such that each element in the matrix can be defined as 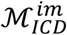.

The ICD-10 catalog contains more than 140000 labels, which is rather intractable in a single model. Since we are dealing with chronic patients, the specific combination of morbidities and co-morbidities imply a smaller ICD space, consisting of 252 items.

This definition implies that each row in 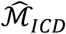 is the corresponding ICD vector of patient *i*, which has a length *m*, and where each element is the number of times an ICD is assigned to each patient, considering that we evaluate the contained data in a time period (for instance 6 months). An example of the 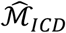 is shown in table 2.

**Table 2.** Exemplary ICD multi-label matrix for the patient population ℳ_ICD_, where the columns are the ICDs and the rows are the patients

|  | Q60.0 | E83.4 | N18.2 | N18.82 | D64.8 | N25.8 | N11.9 | I11.00 | N18.5 | E83.38 | ... | F10.2 | D52.9 | M62.89 | R55 | E14.90 | K27.9 | I44.2 | R73.9 | I80.9 | U80.0 |
| --- | --- | --- | --- | --- | --- | --- | --- | --- | --- | --- | --- | --- | --- | --- | --- | --- | --- | --- | --- | --- | --- |
| 26536 | 41 | 41 | 41 | 41 | 0 | 0 | 0 | 0 | 0 | 0 | ... | 0 | 0 | 0 | 0 | 0 | 0 | 0 | 0 | 0 | 0 |
| 15328 | 0 | 82 | 82 | 0 | 0 | 0 | 0 | 0 | 0 | 0 | ... | 0 | 0 | 0 | 0 | 0 | 0 | 0 | 0 | 0 | 0 |
| 22045 | 0 | 0 | 82 | 0 | 41 | 41 | 0 | 0 | 0 | 0 | ... | 0 | 0 | 0 | 0 | 0 | 0 | 0 | 0 | 0 | 0 |
| 3931 | 0 | 41 | 41 | 41 | 41 | 0 | 0 | 0 | 0 | 0 | ... | 0 | 0 | 0 | 0 | 0 | 0 | 0 | 0 | 0 | 0 |
| 9135 | 41 | 0 | 0 | 0 | 0 | 82 | 41 | 0 | 0 | 0 | ... | 0 | 0 | 0 | 0 | 0 | 0 | 0 | 0 | 0 | 0 |
| ... | ... | ... | ... | ... | ... | ... | ... | ... | ... | ... | ... | ... | ... | ... | ... | ... | ... | ... | ... | ... | ... |
| 26042 | 0 | 0 | 0 | 0 | 0 | 2 | 0 | 0 | 1 | 0 | ... | 0 | 0 | 0 | 0 | 2 | 0 | 0 | 1 | 1 | 1 |
| 3622 | 0 | 0 | 0 | 0 | 0 | 1 | 0 | 0 | 1 | 0 | ... | 0 | 0 | 0 | 0 | 1 | 1 | 4 | 1 | 0 | 1 |
| 22119 | 0 | 0 | 0 | 0 | 0 | 2 | 0 | 0 | 0 | 0 | ... | 0 | 0 | 0 | 0 | 1 | 1 | 0 | 0 | 0 | 2 |
| 19855 | 0 | 0 | 0 | 0 | 0 | 0 | 0 | 0 | 1 | 0 | ... | 0 | 0 | 0 | 0 | 0 | 0 | 1 | 2 | 1 | 3 |
| 13913 | 0 | 0 | 0 | 0 | 0 | 1 | 0 | 0 | 1 | 2 | ... | 0 | 0 | 0 | 0 | 0 | 0 | 0 | 2 | 0 | 1 |

In a similar way, a multi-label matrix of TKs, 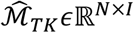 with *N* the number of unique TKs (50 in our study), can also be assigned to the patient’s population, where each row is a vector of length *I* assigned to each patient *i* of unique therapy keys TKs. However, each element in this vector is binary, i.e., the patient gets a given therapy (*ℳ*_*TK*_= 1) or not (*ℳ*_*TK*_= 0). Thus, this definition implies that we essentially have a multilabel problem.

According to this definition, 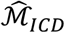 and 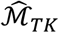 are high dimensional matrices that are difficult to reduce. The heterogeneity of the distribution of the use-frequency of ICDs and TKs poses a challenge: since we are evaluating patients with a characteristic chronic disease associated to specific co-morbidities, there are frequent ICDs and TK combination associated to the more frequent morbidities/co-morbidities, while other diseases are less represented in the total ICD distribution. For this reason, we obtain a heterogeneous ICD distribution (see figure 1).

**Figure 1.**
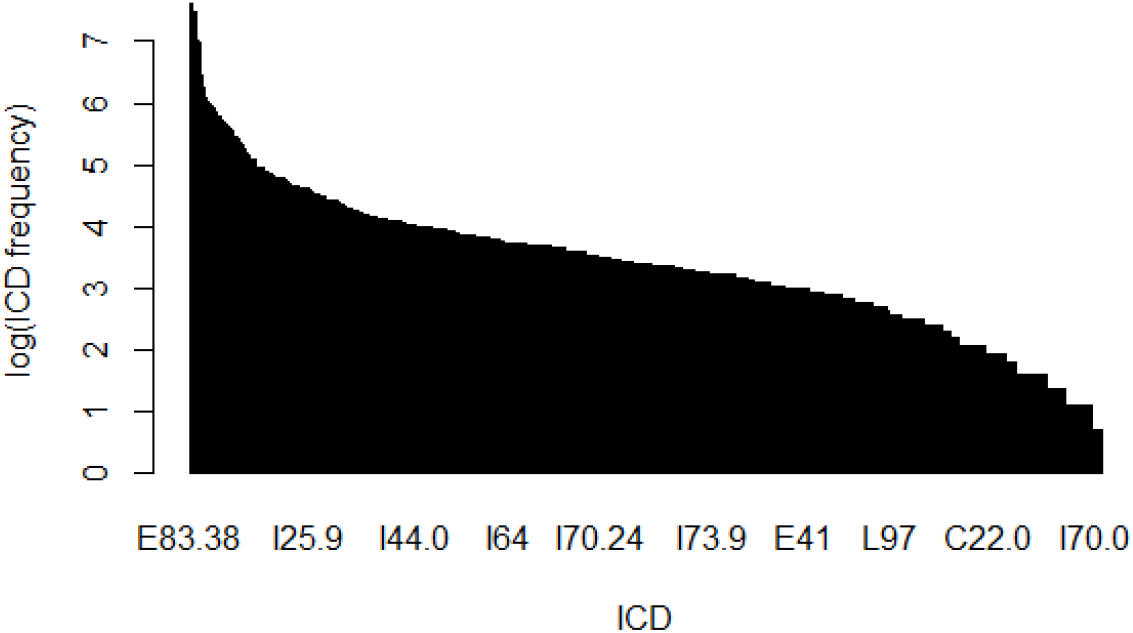
Typical distribution of ICDs, where the y axis is the ICD frequency (as a logarithmic scale). Observe that the distribution is fast scale free, with certain ICDs owning a larger frequency (frequent morbidities) joined by ICDs with less frequency (co-morbidities)

One option to reduce this dimensionality is to cluster the ICDs and TKs. From these clusters we obtain information about the structure of the population depending on the current epidemiological situation (ICD clustering) as well as the medical management of the population (ICD clustering constrained to TKs). However, any clustering methodology implies an ad-hoc way to compress these matrices, which is a procedure where information can get lost. For this reason, we have implemented an autoencoder, which was defined as a feedforward, non-recurrent neural network for dimensionality reduction, such that we transform the 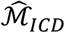 matrix into a new low dimensional matrix 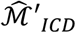, i.e.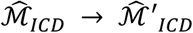.

The input to autoencoder model is standard normalized, i.e., the mean has been removed and the matrix has been scaled to unit variance^4^

This kind of automatic clustering is then used as the input, together with the additional meta parameters containing the patient’s age and gender, for both the baseline models as well as for the GraphNNs. The meta parameters are additionally normalized for the further estimation of the scoring for patient embedding in the matrix: the age of each patient is defined as 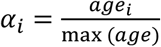, and the binary parameter *µ*_*i*_ is a binary parameter representing the patient’s gender (0 for women, 1 for men).

### 2.3 Baseline models

The combined output of the autoencoder, applied to the 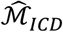 matrix, and the patient’s meta parameters is used as an input feature to train different models aiming to predict the TK labels, i.e., the 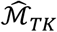 matrix.

The following methods were implemented as baseline models in the current study:

- Random Forest classification (RF)
- Logistic Regression (LR)
- Extra Trees Classifier (ET)
- K-Neighbors Classifier (KNN)
- Decision Tree classifier (DT)

While these methods can be applied in a straightforward way, they strongly compress information about the structure and relatedness between patients, which is important for further clustering analysis. In the next section we demonstrate how Graph-NNs can preserve this information, helping to perform clustering analysis on the patient population. All these models are trained with the default parameters as defined in the SCIKIT-learn library.

### 2.4 Graph clustering

The aim of this method is to perform patient clustering assuming that each patient *k*, owning the same or similar ICD pattern, is linked to other patients with similar ICDs, such that patients with a similar combination of morbidities and comorbidities should get similar TKs. The advantage of this method over traditional clustering is its flexibility, such that new graphs with different connectivity can be easily evaluated.

With this method we do not require ad-hoc definitions or parameters, like the number of clusters by k or c-means clustering methods, since the graph-based clustering establish the direct correlation between ICDs and TKs

*Although embedding methods have proven successful in numerous applications, they suffer from a fundamental limitation: their ability to model complex patterns is inherently bounded by the dimensionality of the embedding space* (Nickel and Kiela, 2017). We solve in this work this problem by performing a patient embedding in a graph object *G*, as is shown in figure 2, such that we encode the corresponding ICD distributions by an object representing an interlinking between similar patients, an object that can be defined as a similarity score 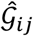, which is essentially the adjacency matrix for the linking between the patient *i* and *j*.

**Figure 2.**
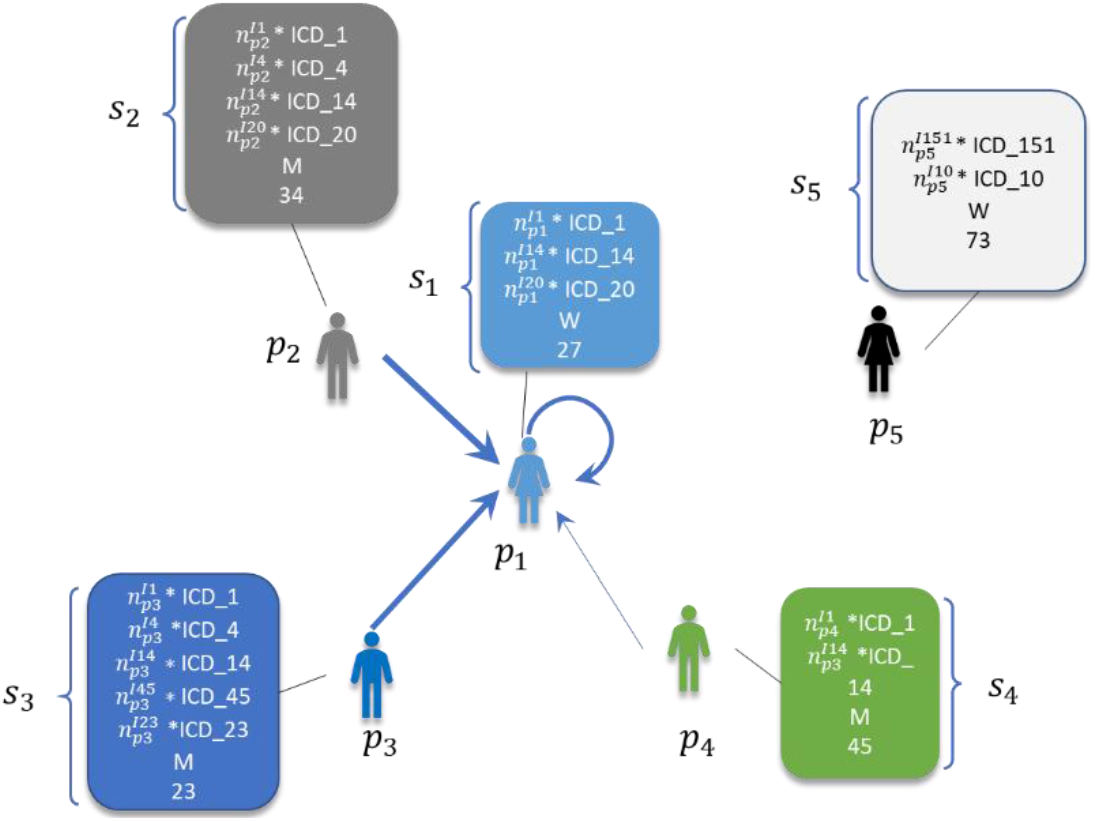
Graph patient embedding

Since a fix clustering method aggregates information in the defined cluster, with graph structures we are aggregating information on each node informing how is the similarity between patients (similarity score (Rocheteau et al., 2021)). This similarity is defined by means of a score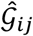, which is a function that depends on the ICD distribution as well as the gender and age relatedness between patient *i* and patient *j*

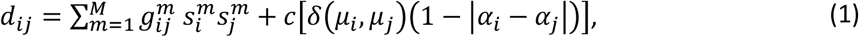

where 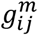 is essentially a weight for the ICD distribution for patients *i* and *j, m* is the number of elements of each of the 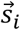 vector, i.e. 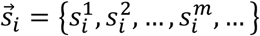, and *M* is the total number of columns of the 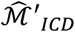 (matrix with reduced dimension from 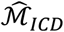 matrix). Additionally, we introduce a scoring that depends on the patient’s meta parameters, where *δ*(*µ*_*i*_, *µ*_*j*_) is the Kronecker’s delta (1 if *µ*_*i*_ = *µ*_*j*_, 0 otherwise), and *c*is a parameter (we call this parameter “importance hyper parameter”) such that patients with a similar gender and similar age get a high score, biasing in this way the initial ICD scoring. The goal of this definition is to bias the graph construction to similar patients based not only on similar diagnoses, but also similar age and gender.

This score will be used to define the strength of the linking between two nodes. The corresponding mathematical structure of the similarity score 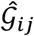 is a function that depends on this relatedness *d*_*ij*_ and the clustering of similar elements in the graph, and which essentially works as the adjacency matrix linking the nodes *p*_*i*_ and *p*_*j*_.

For the definition of this matrix, neighbor elements should be recognized depending on the distance between *p*_*i*_ and *p*_*j*_. For this *computation we estimate the nearest neighbors using a k-Nearest neighbors’ method by finding a predefined number of training samples closest in distance to the new point and predict the label from these* ^5,6^(Altman, 1992), such that

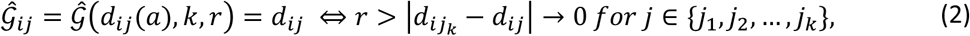

where *k* is the maximal number of nearest neighbors explored by the clustering algorithm, *a* is an internal calibration parameter of the metric, and *r* is the radius where the clustering is performed.

For this evaluation different plausible definitions of the metric can be found. This selection depends on the topology where the points are embedded: since we are dealing with high dimensional spaces with a complex distribution, it is to expect that the best suited metric be non-Euclidean (Bronstein et al., 2017). In our model we define 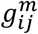 in relation to the total weight of ICDs measured as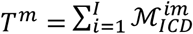, such that one plausible definition of 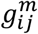 is 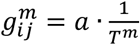, with *a* defined as a constant. Therefore, we give more weight to the ICDs which have low frequency and can be easily neglected during training. Since this definition influences the final definition of the distance in the nearest-neighbors algorithm, we can thus consider 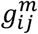 as a definition of the background metric used in the construction of the network. With this modeling strategy we aim to define networks with uniform topologies and avoid the formation of hubs.

Considering this aspect, it is necessary to examine which form the of 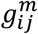 is the more appropriate to perform network encoding. We tested different definitions following the results reported by Nickel and Kiela (Nickel and Kiela, 2017).

The graph encoding, as shown in figure 3, is then used as input for a convolutional neural graph in order to identify corresponding labels for each patient, located in each node of the network^7^. In our case, the labels are the corresponding TKs assigned to each node (patient).

**Figure 3.**
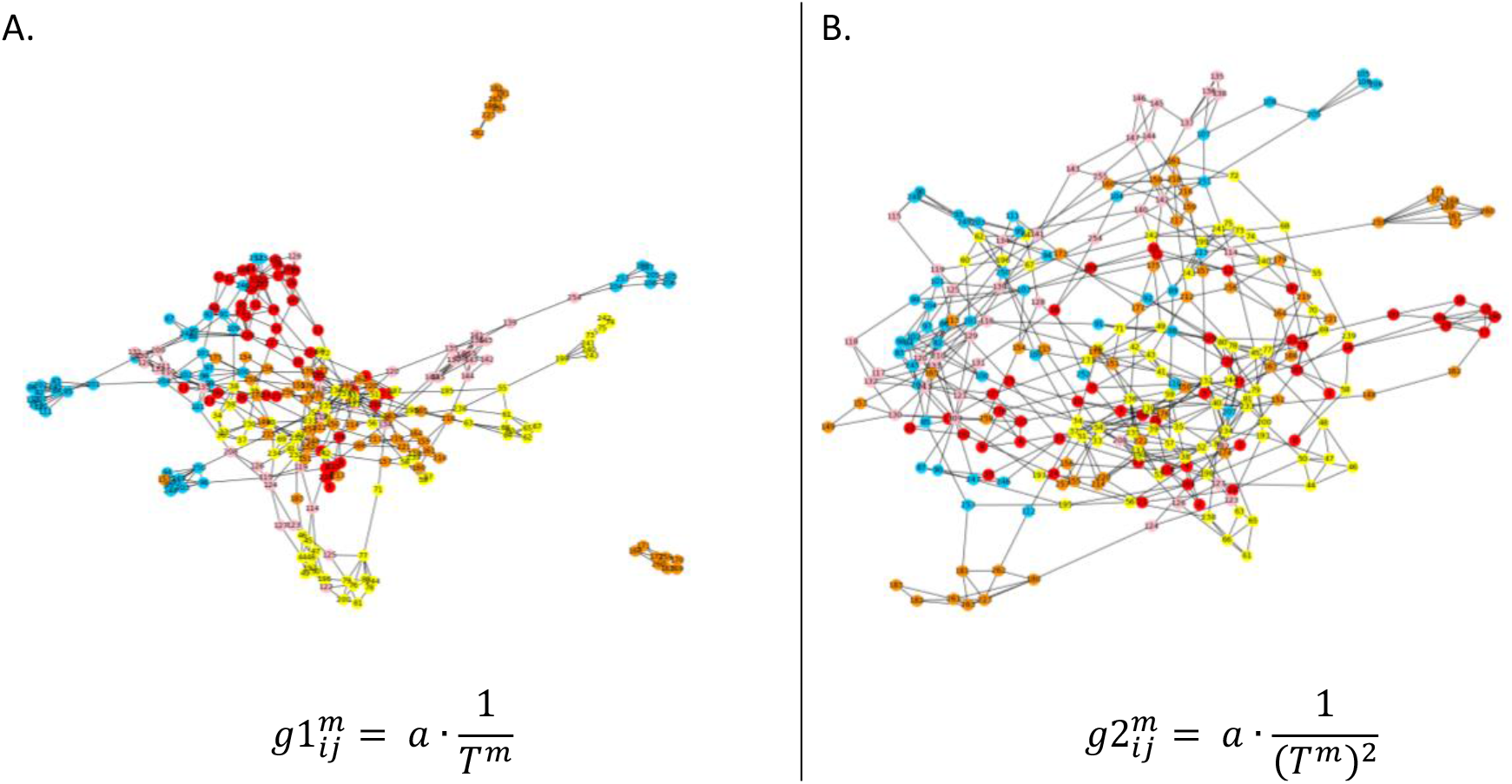
Qualitative comparison of the network encoding linking patients owning similar ICDs, considering two different metric definitions for the computation of patient neighbors. Observe the change in the hub distribution

Since we are considering cumulated events after several periods of time, we essentially encode this information as static embeddings^8^. Additionally, the corresponding model meta parameters are tuned during the model training in short epoch intervals to minimize the training loss and simultaneously get the optimal parameter. The final model structure is provided in the table below.

The labels are the vectors 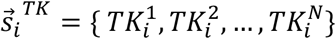, in this case the corresponding TKs at the patient node *i*, with *N* the maximal number of labels, i.e., of TKs. Thus, the trained model M_*GNN*_ takes as input the 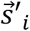 vector containing the corresponding ICDs and meta-parameters of patient *i* and delivers as output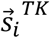, i.e.

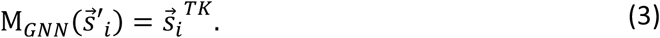

The embedding at the output of the Graph model can be used to make new edges between different patients. This new graph may give us improved edge connections as compared to the graph we build using relatedness score. In order to make a possible edge between node *i* with node *j*, we can use dot product between the node embedding vectors of node *i* and *j*. The dot product score 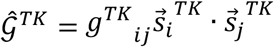 provides is used for the definition of the final adjacency matrix restricted to the TKs, where 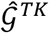 is the similarity score for the TKs. In this case we simply considered that *g*^*TK*^_*ij*_ = 1 (see fig. 4 for the representation of the reconstruction of the TK adjacency matrix).

**Figure 4.**
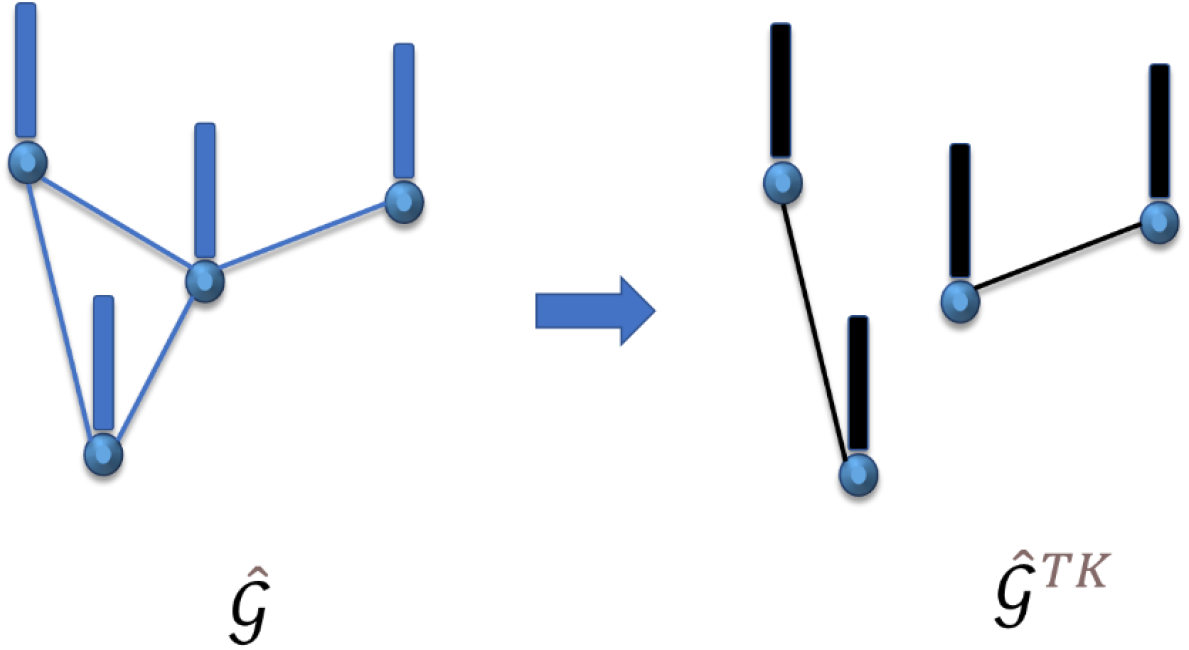
By using the GOPs as labels we are aiming to use an ICD-graph (in blue left) to train a model and predict TKs ordered as a graph as well (in black, right)

The complete workflow, from data collection and low-dimensional data representation using methods for dimension reduction, passing over graph encoding and graph neural networks, to ICD clustering is presented in figure 5. This model has been implemented in Python using Pytorch, with the corresponding libraries for graph neural networks (GNNs).

**Figure 5.**
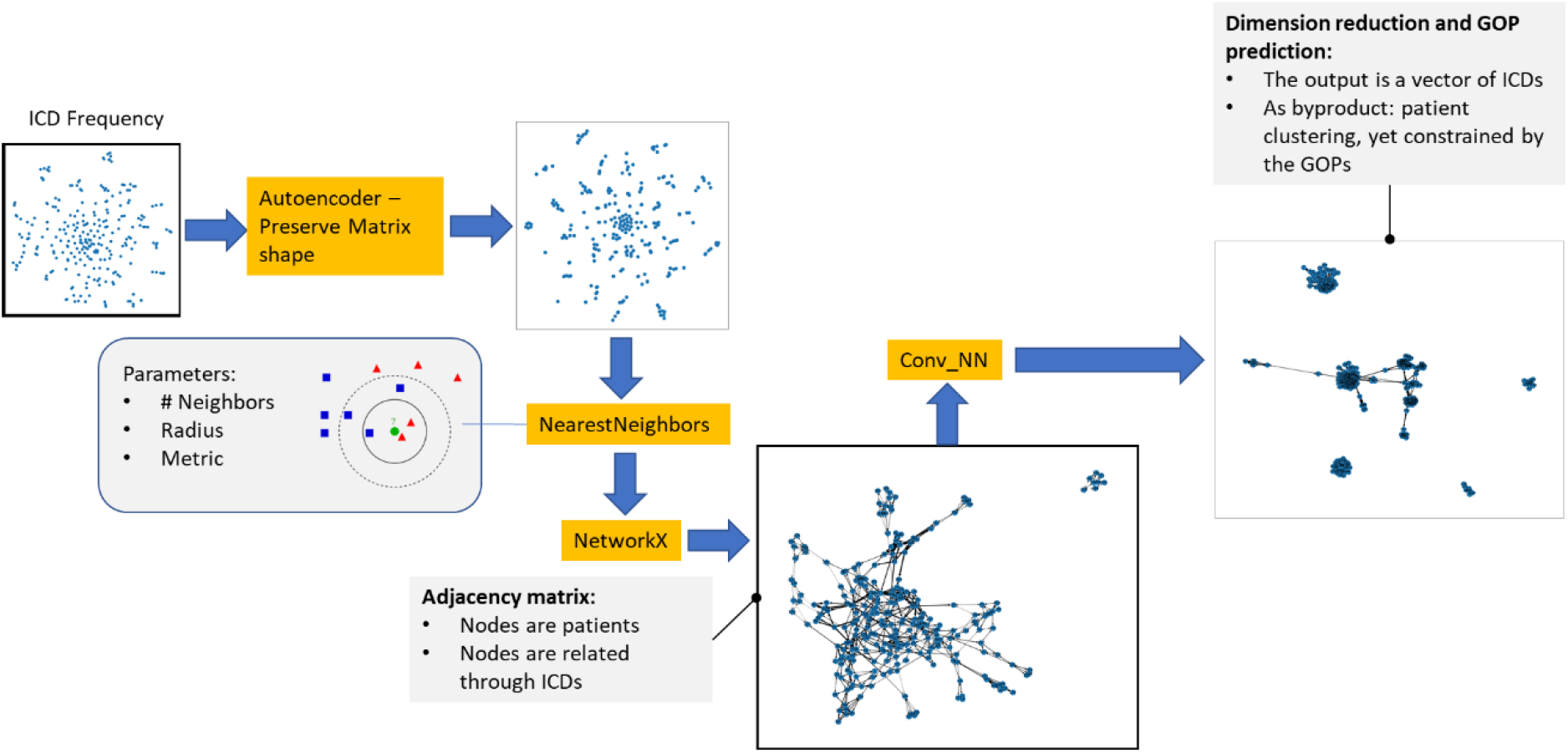
Algorithm’s architecture

In the next section we will present the main results of this study, in particular focusing on the comparison between classical and graph-based clustering methods.

## 3 RESULTS

### 3.1 Validation metrics

The first main goal is to perform a comparative study of the different modelling methodologies. To this end each model is trained using TKs as labels, i.e., the *ℳ*_*TK*_ matrix, implying that the model is essentially a multi-label classification problem for different λ labels. The micro precision was defined as a micro-average

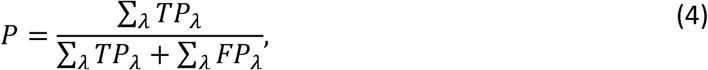

where *TP*_*λ*_ is the total number of observed true positives, and *FP*_*λ*_ the total number of observed false positives of the label λ. Similarly, the micro-recall is defined as

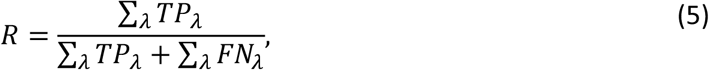

where *FN*_*λ*_ the total number of observed false negatives of the label λ. According to these both definitions, the *f1* score was defined as

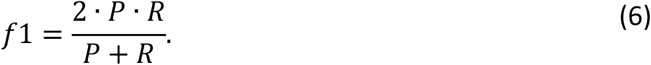

This methodology seems to be plausible considering that in the database we have labels with more instances, and since we want to bias the validation metric towards the most populated ones, we then implemented this kind of micro averaging^9^ in order to bias the final result by class frequency,

### 3.2 Selection OF BESTGNN MODEL: Analysis OF 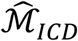 Matrix ON THE Graph Construction

For the first analysis of the graph construction, we focus on the role of the 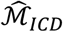 on the graph definition 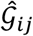, i.e., we assume that *c* = 0. Here, we have performed different validations using models with different 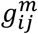 parameters, according to the sore definition in equation 1 and figure 3. The evaluation of the k-Neighbors was made by setting *k* = 3, *a* = 1, and *r* = 1.

We selected these parameters because the results are degraded when the value of *k* other than 3 is used. In particular, larger k values imply an increase of the entropy of the system, which leads to more noise in the final result. Furthermore, we have observed that the radius r has negligible effect on the metric score.

After performing different experiments, we obtained the following validation results for 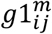 and 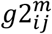, which corresponds to the qualitative results deployed in figure 3 (see table 4). These results imply that there is in fact a dependence on the background geometry where the network is embedded, with a slight influence on the network’s hub distribution.

**Table 3.** Parameters for each layer defined in out GNN model

| Parameter | Meaning | Value | Activation function |
| --- | --- | --- | --- |
| Layer 1 | Convolutional | (252, 32) | relu, dropout 0.5 |
| Layer 2 | Convolutional | (32, 32) | relu, dropout 0.5 |
| Layer 3 | Convolutional | (32, 50) | sigmoid |

**Table 4.** Parameters employed in the K-Nearest Neighbor algorithm

| Weighting | Precision (%) | Recall (%) | f1 (%) |
| --- | --- | --- | --- |
| $g_{ij}^{1m}$ | 86.1 | 69.8 | 77.1 |
| $g_{ij}^{2m}$ | 87.8 | 66.5 | 76.0 |

Furthermore, these parameters have influence on the rank of the scoring parameter 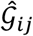 : while 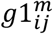 delivers a scaling with values as [0,300], 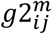 delivers a scaling with values as [0, 8.75] with most values between 0 and 1, which allows a simple coupling to the patient meta parameters. Thus, the following results and final validation were computed using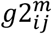.

### 3.3 Final Validation AND Comparison Between Baseline Models and GNN

In the final validation we considered the patient’s meta parameters, i.e., *c*>0. As expected, the coupling to the meta parameters influences the final validation: for *c* = 0.2 we obtained *f*1 = 79.7 (in %), while for *c* = 1.0 we obtained *f*1 = 73.1. For this assessment we would be tempted to select *c*in order to get the best possible validation value. However, this could bias the model too much to the ICD selection and ignore the fact that the TK selection depends on the patient’s age and gender as well. Since the scaling of the selected metric has the most values between 0 and 1, we fixed *c* = 1.0, which delivers a rank scoring 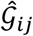 of [0,2] in order to give importance to the patient’s meta parameters.

In this case we selected the best performing GNN model and compared it against the baseline models to assess the quality and eventual superiority of the GNN models. The results of this comparison are resumed in the table 5 (observe that the scores for the GNN models are smaller since we are considering here the patient’s meta parameters).

**Table 5.**
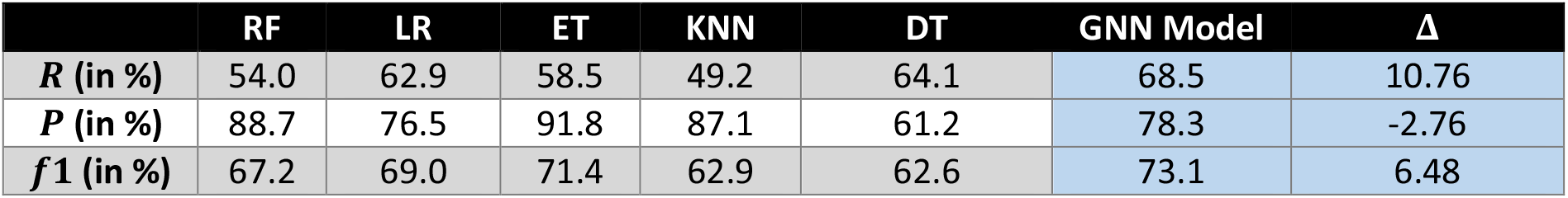
Comparative validation results between baseline models and GNNs

| | RF | LR | ET | KNN | DT | GNN Model | $\Delta$ |
| --- | --- | --- | --- | --- | --- | --- | --- |
| **R (in %)** | 54.0 | 62.9 | 58.5 | 49.2 | 64.1 | 68.5 | 10.76 |
| **P (in %)** | 88.7 | 76.5 | 91.8 | 87.1 | 61.2 | 78.3 | -2.76 |
| **f1 (in %)** | 67.2 | 69.0 | 71.4 | 62.9 | 62.6 | 73.1 | 6.48 |

From the method comparison we can observe that the Graph-NN provides satisfactory results regarding the other methods. After testing the GNN model over test data we obtained a *f1* score of 73.1%, which is satisfactory regarding the baseline models; only RF was superior in precision than the GNN. These results suggest that GNNs seems to outperform the other methods, with an average difference of 10.76 for recall, -2.76 for precision, and 6.48 for *f1*. Furthermore, this methodology allows a detailed clustering analysis of the patient population, helping to better interpret results and obtain further relevant information in the population.

### 3.4 Clustering analysis of TKS

Based on 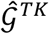 we are able to reconstruct a new network interlinking the patients depending on the TKs. This new network reveals a structure with clustered patients, such that each cluster reveals the similitude between the patients depending on their therapies.

This implies, from the initial ICD-patient network 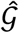 we obtain a structure that defines how similar are the TKs assigned to individual patients.

Since we have different health centers, we have hypothesized that this clustering is directly correlated to the health center distribution (geographical clustering represented by different colors, see figure 6). However, the assignation of the 5 different health centers to the clusters reveals that such clusters originate from similar patient therapies, and not from the geographic distribution. Thus, with this modelling we are able to identify how patients are usually treated, helping in this way to perform an empirical identification of specific medical guidelines used to treat the patient populations.

**Figure 6.**
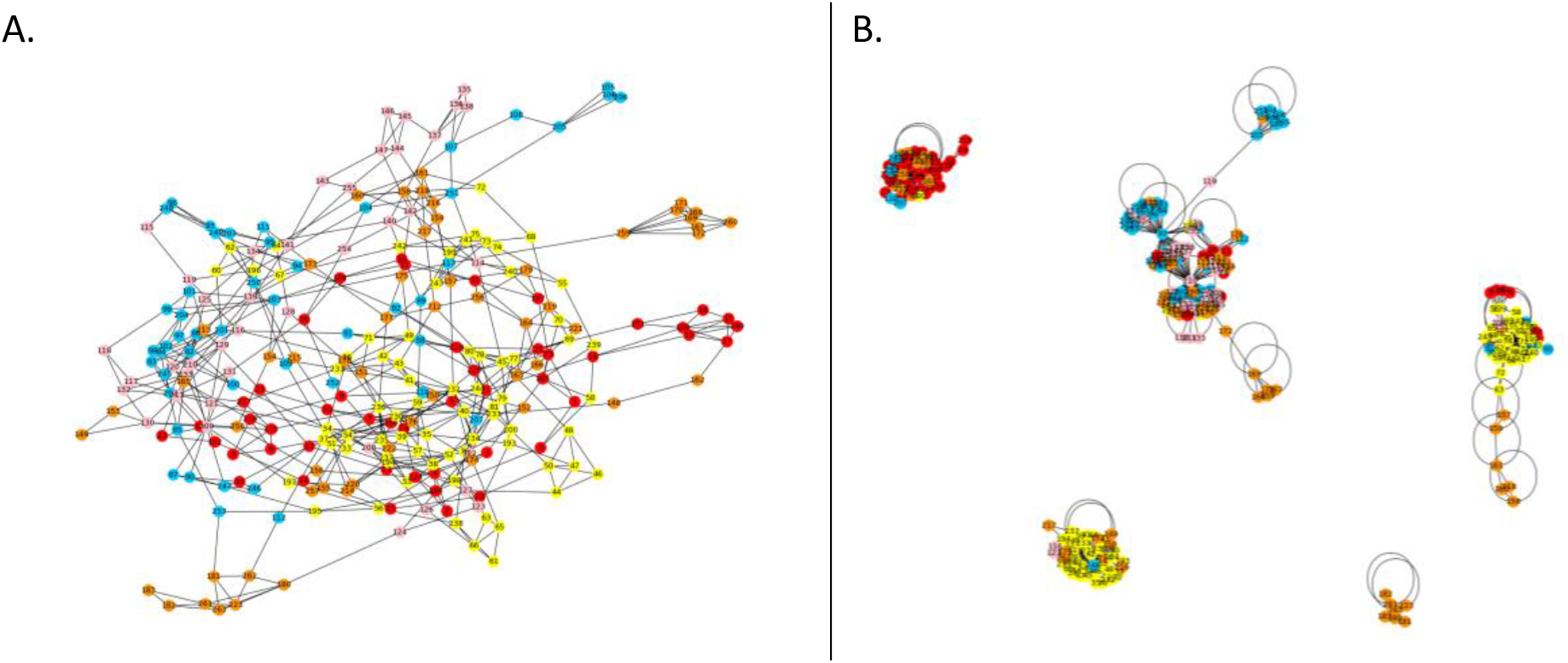
Graph patient structure defined on the assignation of similar TKs between patients. Left, input ICD graph, right output TK graph. Different colors correspond to different health centers (geographical distribution)

Since these guidelines are correlated to the ICDs, we analyze the relation between the ICDs (left graph structure in figure 6) and the TKs (right graph structure in figure 6) by measuring the difference between both structures. To this end we employ a saliency method. This method, often used in natural language processing (NLP) and image recognition, allows the analysis of the significance of input features for the computation of the output. Thus, in language recognition this implies for instance the recognition of relevant words used for the composition of sentences (Samardzhiev et al., 2017), or in image recognition the pixel ranking leading to an specific class classification for an image (Simonyan et al., 2014), in order to understand how the model is looking into the image^10^.

In this investigation we define the saliency score in the same way as Simonyan et al. (Simonyan et al., 2014), where the score of a label *Sc*_*TK*_ is in principle a non-linear function that can be approximated by a Taylor approximation to a linear function to the model input, in this case the ICD matrix, such that 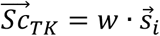, where *w* is the saliency (or importance) score. The score *w*can in this case be computed by back propagation, i.e., the gradient of the model output (difference of number of times a single label is observed) respect to the input (different single combinations of ICDs) 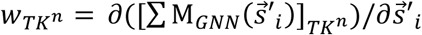 (from equation 3), which depend on the vector 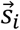, i.e., for *TK*^*n*^ then 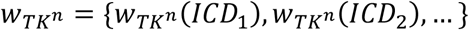. This score is computed either for the whole label-network *w*_*all*_ as well as for each one of the sub-networks *w*_*c*_, in this case 5 subnetworks according to figure 6 B.

In this way, the saliency score basically measures the “prominence” of an ICD for the definition of a TK based on the analysis of the whole population.

In Fig. 7 we deploy the saliency score (y axis), where we have basically discovered 5 different ICD groups for each TK cluster (clusters represented in figure 6 B). By means of this procedure we can thus envision a pyramid structure, where specific patient clusters are related to specific ICDs and GOPs, leading to the discovery of specific guidelines of certain co-morbidities distributed around a main morbidity (ICD with highest distribution in figure 7). The main question to answer here is “which ICDs are important for predicting TKs? “. The x axis of the plots is the ICDs that are important for the prediction of TKs for the graphs/ subgraphs. The y axis shows how the importance of those ICDs is based on the saliency score. Last plot shows the importance of different ICDs for the complete graph.

**Figure 7.**
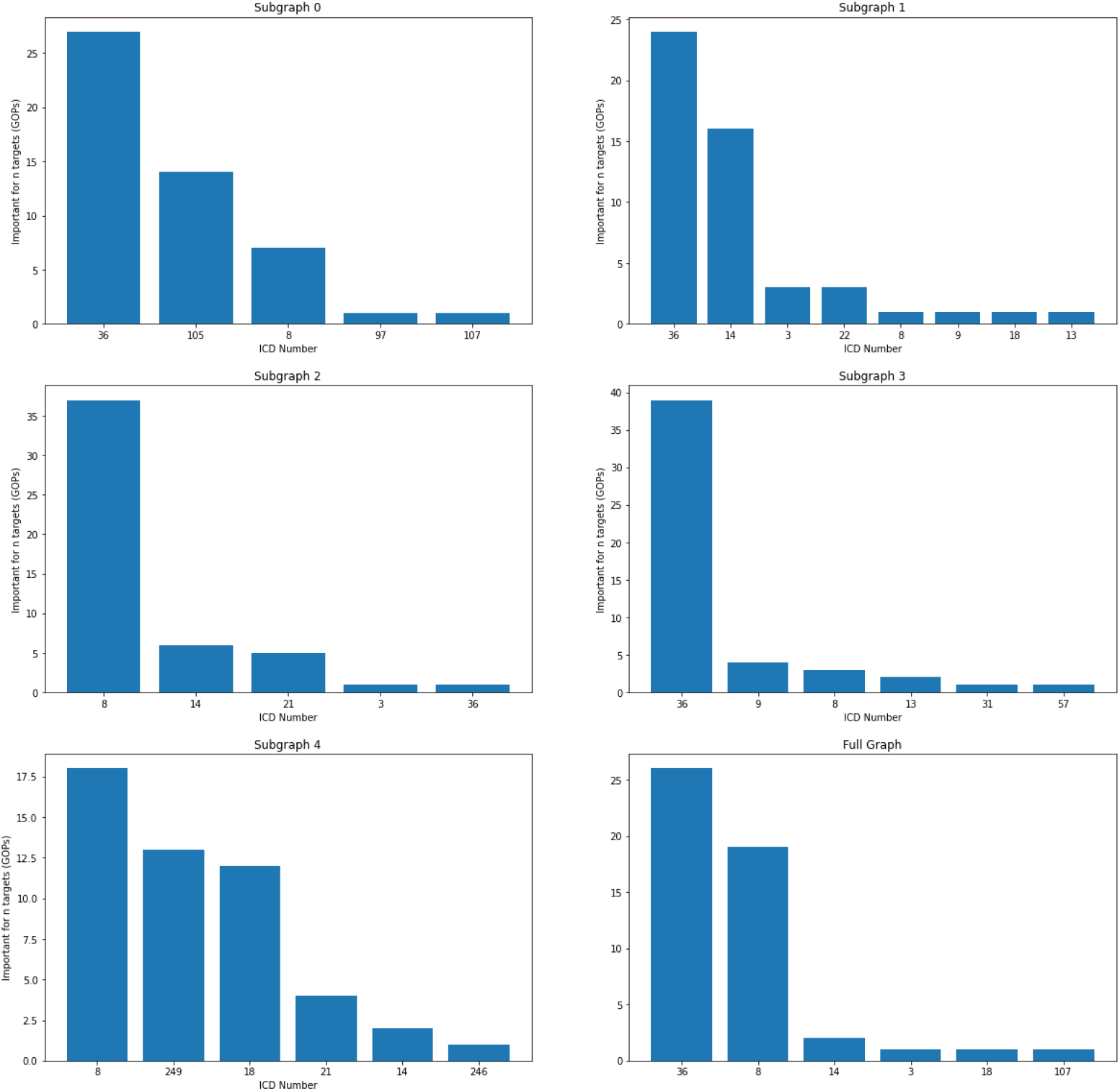
Cluster of main morbidities (ICD with high distribution, for instance ICD number 5) combined to co-morbidities leading into specific combination of TKs (clusters in figure 6)

Each plot of subgraph also contains ICDs present in full graph. However, in addition to these ICDs that are important for the complete graph, each subgraph have additional ICDs that are important for that subgraph e.g for subgraph 0, ICD numbers 36 and 105 are important. However, the ICD number 105 does not seem to be important in the full graph, i.e., for the whole population. This demonstrates that clusters are probably made based on different TKs; and for predicting those TKs different types of ICDs are hence required.

## 4 Discussion

In this work, we conducted a comparison between different AI methods and GNNs to extract patterns from EHRs to empirically derive guidelines currently used in a healthcare system. To this end, we have implemented conventional modeling techniques in AI/ML to discover ICD-TK correlations, which are the product of implicit guidelines currently used by the medical community.

While classical AI/ML approaches and GNNs provide similar validation results, we have discovered that GNNs can both provide better validation results and identify clusters where certain combinations of morbidities/comorbidities are related to specific TK combinations. Therefore, GNNs allows us to perform additional cluster analyses that are difficult to perform with other methods due to their inherent lack of internal structure. This confirms other findings from other groups that have highlighted the advantage of graph methods for the analysis of health data.(Kotwal et al., 2016).

Of course, this methodology is not applicable to all 140000 ICD-10 labels, but a tool that is better suited for dealing with patients with specific morbidities. This implies that before applying this methodology, a dimension reduction of the ICD landscape is required.

Despite the advantage in using graphs to both predict and provide structures leading to cluster analysis, we are aware of the problem of using networks paradigm due that they can introduce undesired bias: in several fields we observe how network-methods sometimes establish incorrect interlinking between elements that are not truly correlated, for instance the familiar linking of an individual to a criminal family does not imply that the individual is a criminal. Network paradigms reinforce biases and can be even dangerous, in particular when indicators are used to encode features in graph structures (Sugimoto, 2021), in particular when these network representation is viewed as a fundamental natural characteristic of social and biological systems. In this project we observe the same problem, since Graph information encoding (GIE) based only on ICDs might ignore relevant medical information encoded on other sources, like physician notes or sentiment analysis.

One option to solve these problems is to continue making data enrichment to the present structures including additional data sources. However, these methods do not avoid the risk to bias the model by stablishing links, and more research is required to avoid eventual misleading conclusions. Furthermore, the ICDs self are not free of error: according to some studies, only 63% of the ICDs is accurate^11^, which implies that the inputs used in the model might not be accurately enough.

An additional observation concerns the graph construction and the focus on validation score of the model: a model with a small coupling of the patient`s meta parameters with ICDs leads to a better validation of the model, with a score *f1* of 77%, which is much better than the reported score of 73%. However, this could give less relevance to factors like gender and age to the modeling. But both parameters are actually very relevant to avoid model bias, for instance considering that the ICD combination for a woman can lead to a different therapy than for a man. Therefore, this is an example of how it is better to accept lower accuracy in order to preserve and represent basic characteristics of the population.

Finally, the databases are evolving, implying that derived network structures are not valid for different times. To this end, and following the work of Rocheteau et al. (Rocheteau et al., 2021) we plan to implement methods to consider time series and couple this information into the node characterization according to figure 4^12^.

## 5 Conclusion

We have reported the use of GNNs for the identification of correlation patterns between ICDs and TKs. We have demonstrated that this modelling method has a high accuracy respect base-line models, since it allows a more accurate identification of patient clusters based on their ICDs as other similar methods. Furthermore, this methodology allows a better identification of patient groups with specific TK groups correlated to specific ICD combinations. This result is therefore helpful to identify pyramid-like structures in the ICD distribution, which is an important information not only to leverage the quality of the patient management (depending on specific ICD combinations) but also to better identify the performance of health centers by region.

Despite these positive results, an improvement in the ICD identification as well as better methods to reduce potential bias induced by the network models is required to further improve this technology. Furthermore, tests should be performed directly on EHRs in order to corroborate the advantage of this methodology using authentic data before using it as an interventional method.

## Data Availability

All data produced in the present study are available upon reasonable request to the authors

## Data availability and ethical issues

The data used in this study is fully synthetic. All data produced in the present study are available upon reasonable request to the authors

## Acknowledgments

We are very grateful with Martin Grundman and Thomas Shimper for relevant inputs and ideas in the formulation of the motivation and applicability of this methodology for the analysis of health records, and Felix Weil for his constant support for the development of this work.

## Author contribution

JGDO and FM conceived the idea; JGDO implemented and performed the data synthetization; FM implemented the GNNs routine and wrote the python notebook; JGDO performed further modifications on the notebook, conceived the artwork and wrote the first draft of the article. FM and JGDO wrote together the final article version.

## Abbreviations

GNN: Graph Neural Networks
HER: Electronic Health Record
PHI: Patient Health Information
IQI: Inpatient Quality Indicators
PQI: Prevention Quality Indicators
PSI: Patient Safety Indicators
TK: Therapy Keys (equivalent to medical procedures)
ICD: International Classification of Diseases
ID: Internal Patient identification

## Footnotes

1 for example, of COVID-19 patients

2 https://github.com/NYUMedML/GNN_for_EHR

3 https://cmte.ieee.org/futuredirections/tech-policy-ethics/2018articles/ethical-issues-in-secondary-use-of-personal-health-information/

4 https://scikit-learn.org/stable/modules/generated/sklearn.preprocessing.StandardScaler.html

5 https://scikit-learn.org/stable/modules/neighbors.html

6 [NearestNeighbors](https://scikit-learn.org/stable/modules/generated/sklearn.neighbors.NearestNeighbors.html#sklearn.neighbors.NearestNeighbors) implements unsupervised nearest neighbors learning. It acts as a uniform interface to three different nearest neighbors algorithms: [BallTree](https://scikit-learn.org/stable/modules/generated/sklearn.neighbors.BallTree.html#sklearn.neighbors.BallTree), [KDTree](https://scikit-learn.org/stable/modules/generated/sklearn.neighbors.KDTree.html#sklearn.neighbors.KDTree), and a brute-force algorithm based on routines in sklearn.metrics.pairwise.

7 See table 4 for the final validation results and final assessment of the best possible weight 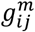

8 In future applications we must consider the evolution of graphs according to changes in the graph-encoding in different time periods, which must lead to a dynamic network structure.

9 https://stats.stackexchange.com/questions/156923/should-i-make-decisions-based-on-micro-averaged-or-macro-averaged-evaluation-mea

10 And for this reason this method is used for AI explainability. See e.g. https://medium.datadriveninvestor.com/visualizing-neural-networks-using-saliency-maps-in-pytorch-289d8e244ab4

11 https://healthinfoservice.com/most-common-icd-10-error-codes/

12 For instance, using LSTM methods to consider events from different times as an additional node feature

